## Supplemental Information for "Social disparities in unplanned 30-day readmission rates after hospital discharge in patients with chronic health conditions: A retrospective cohort study using patient level hospital administrative data linked to the population census in Switzerland" (from another version of this preprint: version 1, 2022-01-21)

**Supporting Information**

**S1 Table. Odds ratios of fully adjusted logistic regression for risk of unplanned 30-day readmission (all-conditions model C) with interaction chronic condition*education level (N=62,109)**

| **Outcome: risk for 30-day readmission** | **Model C with CHC*education level** | | | |
| --- | --- | --- | --- | --- |
|  | Sig. | Exp(B) | 95% CI | |
|  |  |  | Lower | Upper |
| Education level |  |  |  |  |
| tertiary (ref.) | 0.252 |  |  |  |
| upper secondary | 0.194 | 1.3 | 0.875 | 1.933 |
| compulsory | 0.097 | 1.397 | 0.941 | 2.072 |
| Insurance class |  |  |  |  |
| mandatory (ref.) |  |  |  |  |
| (Semi-)private | 0.035 | 0.893 | 0.804 | 0.992 |
| Household type |  |  |  |  |
| Living with others (ref.) |  |  |  |  |
| Living alone | 0.149 | 1.077 | 0.974 | 1.192 |
| Sex |  |  |  |  |
| Men (ref.) |  |  |  |  |
| Women | 0.031 | 0.891 | 0.803 | 0.989 |
| Language skills |  |  |  |  |
| At least regional language or English (ref.) |  |  |  |  |
| Not regional language and no English | 0.41 | 1.06 | 0.923 | 1.217 |
| Age |  |  |  |  |
| <= 55 years | <.001 |  |  |  |
| 56-65 years | 0.001 | 1.318 | 1.116 | 1.556 |
| 67-74 years | <.001 | 1.6 | 1.357 | 1.886 |
| 75+ years | <.001 | 1.98 | 1.686 | 2.326 |
| Chronic health condition |  |  |  |  |
| Ischaemic heart disease (ref.) | <.001 |  |  |  |
| Lung cancer | 0.243 | 1.424 | 0.787 | 2.575 |
| Colon cancer | 0.061 | 0.471 | 0.214 | 1.037 |
| Breast cancer | 0.007 | 0.42 | 0.223 | 0.791 |
| Prostate cancer | 0.989 | 0.997 | 0.627 | 1.584 |
| Diabetes with/without complications | 0.91 | 0.964 | 0.512 | 1.816 |
| Acute myorcardial infarction | 0.01 | 0.503 | 0.298 | 0.849 |
| Acute cerebrovascular diseases | 0.065 | 0.593 | 0.34 | 1.032 |
| Congestive heart failure | <.001 | 0.395 | 0.244 | 0.642 |
| COPD or asthma | 0.067 | 0.494 | 0.232 | 1.051 |
| Osteoarthritis | <.001 | 0.294 | 0.188 | 0.461 |
| Back problems and disc order | <.001 | 0.336 | 0.21 | 0.537 |
| Comorbidity |  |  |  |  |
| NSD, centered by CHC | <.001 | 1.145 | 1.108 | 1.183 |
| Mental comorbidity: no (ref.) |  |  |  |  |
| Mental comorbidity: yes | 0.02 | 1.188 | 1.028 | 1.373 |
| Previous hospital stay last 6 months |  |  |  |  |
| No (ref.) |  |  |  |  |
| Yes | <.001 | 1.771 | 1.587 | 1.976 |
| Length of hospital stay (LOS) |  |  |  |  |
| LOS, centred by CHC, Q1 (ref.) | <.001 |  |  |  |
| LOS, centred by CHC, Q2 | 0.412 | 0.943 | 0.82 | 1.084 |
| LOS, centred by CHC, Q3 | 0.257 | 1.079 | 0.946 | 1.232 |
| LOS, centred by CHC, Q4 | <.001 | 1.637 | 1.445 | 1.856 |
| CHC*Education level |  |  |  |  |
| Ischaemic heart disease (ref.) | 0.074 |  |  |  |
| Lung cancer*upper secondary | 0.213 | 1.516 | 0.788 | 2.919 |
| Lung cancer*compulsory | 0.291 | 1.429 | 0.737 | 2.769 |
| Colon cancer*upper secondary | 0.332 | 1.539 | 0.644 | 3.68 |
| Colon cancer*compulsory | 0.465 | 1.404 | 0.565 | 3.493 |
| Breast cancer*upper secondary | 0.576 | 0.817 | 0.403 | 1.658 |
| Breast cancer*compulsory | 0.743 | 0.886 | 0.429 | 1.829 |
| Prostate cancer*upper secondary | 0.081 | 0.611 | 0.351 | 1.063 |
| Prostate cancer*compulsory | 0.23 | 0.687 | 0.372 | 1.269 |
| Diabetes with/without complications*upper secondary | 0.312 | 0.683 | 0.326 | 1.43 |
| Diabetes with/without complications*compulsory | 0.127 | 0.554 | 0.26 | 1.182 |
| Acute myorcardial infarction*upper secondary | 0.622 | 1.16 | 0.643 | 2.095 |
| Acute myorcardial infarction*compulsory | 0.74 | 0.899 | 0.48 | 1.684 |
| Acute cerebrovascular diseases*upper secondary | 0.816 | 0.927 | 0.491 | 1.75 |
| Acute cerebrovascular diseases*compulsory | 0.671 | 1.148 | 0.607 | 2.173 |
| Congestive heart failure*upper secondary | 0.457 | 0.809 | 0.463 | 1.414 |
| Congestive heart failure*compulsory | 0.946 | 1.02 | 0.578 | 1.8 |
| COPD or asthma*upper secondary | 0.295 | 1.543 | 0.686 | 3.472 |
| COPD or asthma*compulsory | 0.642 | 1.214 | 0.536 | 2.75 |
| Osteoarthritis*upper secondary | 0.036 | 0.574 | 0.343 | 0.963 |
| Osteoarthritis*compulsory | 0.068 | 0.607 | 0.356 | 1.037 |
| Back problems and disc order*upper secondary | 0.447 | 1.225 | 0.726 | 2.066 |
| Back problems and disc order*compulsory | 0.437 | 1.234 | 0.726 | 2.097 |
| Constant | <.001 | 0.032 |  |  |
| Omnibus Chi^2^ | 1580.411(48). <.001 | | | |
| "-2 Log-Likelihood" | 16699.63 | | | |
| ROC | 0.743 | | | |

**S2 Table. Odds ratios of fully adjusted logistic regression for risk of unplanned 30-day readmission (all-condition model C) with interaction chronic condition*household type (N=62,109)**

| **Outcome: risk for 30-day readmission** | **Model C with CHC*household type** | | | |
| --- | --- | --- | --- | --- |
|  | Sig. | Exp(B) | 95% CI | |
|  |  |  | Lower | Upper |
| Education level |  |  |  |  |
| tertiary (ref.) | 0.002 |  |  |  |
| upper secondary | 0.005 | 1.205 | 1.057 | 1.373 |
| compulsory | <.001 | 1.298 | 1.122 | 1.502 |
| Insurance class |  |  |  |  |
| mandatory (ref.) |  |  |  |  |
| (Semi-)private | 0.031 | 0.891 | 0.802 | 0.99 |
| Household type |  |  |  |  |
| Living with others (ref.) |  |  |  |  |
| Living alone | 0.024 | 1.443 | 1.049 | 1.985 |
| Sex |  |  |  |  |
| Men (ref.) |  |  |  |  |
| Women | 0.036 | 0.894 | 0.806 | 0.993 |
| Language skills |  |  |  |  |
| At least regional language or English (ref.) |  |  |  |  |
| Not regional language and no English | 0.423 | 1.058 | 0.921 | 1.215 |
| Age |  |  |  |  |
| <= 55 years | <.001 |  |  |  |
| 56-65 years | 0.001 | 1.329 | 1.125 | 1.57 |
| 67-74 years | <.001 | 1.612 | 1.367 | 1.9 |
| 75+ years | <.001 | 1.988 | 1.692 | 2.336 |
| Chronic health condition |  |  |  |  |
| Ischaemic heart disease (ref.) | <.001 |  |  |  |
| Lung cancer | <.001 | 6.474 | 5.007 | 8.369 |
| Colon cancer | <.001 | 2.076 | 1.485 | 2.901 |
| Breast cancer | 0.335 | 1.169 | 0.851 | 1.606 |
| Prostate cancer | <.001 | 2.111 | 1.619 | 2.753 |
| Diabetes with/without complications | 0.003 | 1.739 | 1.214 | 2.489 |
| Acute myorcardial infarction | <.001 | 1.62 | 1.248 | 2.104 |
| Acute cerebrovascular diseases | <.001 | 1.765 | 1.335 | 2.332 |
| Congestive heart failure | <.001 | 3.007 | 2.368 | 3.82 |
| COPD or asthma | <.001 | 2.148 | 1.611 | 2.866 |
| Osteoarthritis | <.001 | 0.588 | 0.463 | 0.746 |
| Back problems and disc order | 0.078 | 1.228 | 0.977 | 1.544 |
| Comorbidity |  |  |  |  |
| NSD, centred by CHC | <.001 | 1.145 | 1.108 | 1.183 |
| Mental comorbidity: no (ref.) |  |  |  |  |
| Mental comorbidity: yes | 0.018 | 1.191 | 1.03 | 1.376 |
| Previous hospital stay last 6 months |  |  |  |  |
| No (ref.) |  |  |  |  |
| Yes | <.001 | 1.777 | 1.592 | 1.982 |
| Length of hospital stay (LOS) |  |  |  |  |
| LOS, centred by CHC, Q1 (ref.) | <.001 |  |  |  |
| LOS, centred by CHC, Q2 | 0.43 | 0.945 | 0.822 | 1.087 |
| LOS, centred by CHC, Q3 | 0.253 | 1.08 | 0.946 | 1.233 |
| LOS, centred by CHC, Q4 | <.001 | 1.64 | 1.448 | 1.859 |
| CHC*Household type |  |  |  |  |
| Ischaemic heart disease (ref.) | 0.651 |  |  |  |
| Lung cancer*living alone | 0.035 | 0.606 | 0.381 | 0.966 |
| Colon cancer*living alone | 0.202 | 0.659 | 0.348 | 1.25 |
| Breast cancer*living alone | 0.09 | 0.632 | 0.372 | 1.074 |
| Prostate cancer*living alone | 0.581 | 0.861 | 0.507 | 1.464 |
| Diabetes with/without complications*living alone | 0.982 | 0.994 | 0.569 | 1.736 |
| Acute myorcardial infarction*living alone | 0.163 | 0.712 | 0.442 | 1.148 |
| Acute cerebrovascular diseases*living alone | 0.442 | 0.83 | 0.516 | 1.335 |
| Congestive heart failure*living alone | 0.193 | 0.769 | 0.517 | 1.142 |
| COPD or asthma*living alone | 0.071 | 0.647 | 0.403 | 1.038 |
| Osteoarthritis*living alone | 0.127 | 0.71 | 0.456 | 1.103 |
| Back problems and disc order*living alone | 0.108 | 0.721 | 0.484 | 1.074 |
| Constant | <.001 | 0.011 |  |  |
| Omnibus Chi^2^ | 1556.755(37). <.001 | | | |
| "-2 Log-Likelihood" | 16723.28 | | | |
| ROC | 0.741 | | | |

**S3 Table. Odds ratios of multivariate logistic regression for risk of unplanned 30-day readmission by social factors, health status and length of stay in hospital for all heart diseases and diabetes (N total=19,324°/N readmissions=897)**

|  | **A: Social factors** | | | | **B: Health status** | | | | **C: Length of stay** | | | |
| --- | --- | --- | --- | --- | --- | --- | --- | --- | --- | --- | --- | --- |
|  | Sig. | Exp(B) | 95% CI | | Sig. | Exp(B) | 95% CI | | Sig. | Exp(B) | 95% CI | |
|  |  |  | Lower | Upper |  |  | Lower | Upper |  |  | Lower | Upper |
| Education level |  |  |  |  |  |  |  |  |  |  |  |  |
| tertiary (ref.) | 0.005 |  |  |  | 0.013 |  |  |  | 0.018 |  |  |  |
| upper secondary | 0.049 | 1.224 | 1.001 | 1.498 | 0.069 | 1.207 | 0.985 | 1.478 | 0.069 | 1.207 | 0.985 | 1.478 |
| compulsory | 0.001 | 1.435 | 1.151 | 1.789 | 0.004 | 1.39 | 1.114 | 1.735 | 0.005 | 1.374 | 1.101 | 1.715 |
| Insurance class |  |  |  |  |  |  |  |  |  |  |  |  |
| mandatory (ref.) |  |  |  |  |  |  |  |  |  |  |  |  |
| (Semi-)private | 0.053 | 0.854 | 0.727 | 1.002 | 0.095 | 0.872 | 0.742 | 1.024 | 0.14 | 0.885 | 0.753 | 1.041 |
| Household type |  |  |  |  |  |  |  |  |  |  |  |  |
| Living with others (ref.) |  |  |  |  |  |  |  |  |  |  |  |  |
| Living alone | 0.019 | 1.198 | 1.03 | 1.394 | 0.027 | 1.187 | 1.02 | 1.382 | 0.043 | 1.17 | 1.005 | 1.362 |
| Sex |  |  |  |  |  |  |  |  |  |  |  |  |
| Men (ref.) |  |  |  |  |  |  |  |  |  |  |  |  |
| Women | 0.686 | 0.968 | 0.829 | 1.132 | 0.779 | 0.978 | 0.836 | 1.143 | 0.735 | 0.973 | 0.833 | 1.138 |
| Age (years) | <.001 | 1.039 | 1.033 | 1.046 | <.001 | 1.033 | 1.026 | 1.039 | <.001 | 1.031 | 1.025 | 1.038 |
| Comorbidity |  |  |  |  |  |  |  |  |  |  |  |  |
| NSD centred by CHC, below average (ref.)* |  |  |  |  |  |  |  |  |  |  |  |  |
| average |  |  |  |  | 0.584 | 0.946 | 0.775 | 1.155 | 0.459 | 0.927 | 0.759 | 1.133 |
| above average |  |  |  |  | <.001 | 1.403 | 1.182 | 1.666 | 0.003 | 1.296 | 1.089 | 1.542 |
| Mental comorbidity: no (ref.) |  |  |  |  |  |  |  |  |  |  |  |  |
| Mental comorbidity: yes |  |  |  |  | 0.035 | 1.257 | 1.016 | 1.555 | 0.111 | 1.19 | 0.961 | 1.473 |
| Previous hospital stay last 6 months |  |  |  |  |  |  |  |  |  |  |  |  |
| No (ref.) |  |  |  |  |  |  |  |  |  |  |  |  |
| Yes |  |  |  |  | <.001 | 2.16 | 1.851 | 2.521 | <.001 | 2.189 | 1.875 | 2.556 |
| LOS, centred by CHC, Q1-Q3 (Ref.) |  |  |  |  |  |  |  |  |  |  |  |  |
| LOS, centred by CHC, Q4 |  |  |  |  |  |  |  |  | <.001 | 1.643 | 1.418 | 1.903 |
| Constant | <.001 | 0.002 |  |  | <.001 | 0.003 |  |  | <.001 | 0.003 |  |  |
| Omnibus Chi^2^ | 231.29(6) p<.001 | | | | 357.16(10) p<.001 | | | | 399.02(11) p<.001 | | | |
| "-2 log-likelihood" | 7028.08 | | | | 6902.21 | | | | 6860.34 | | | |
| ROC | 0.649 | | | | 0.687 | | | | 0.697 | | | |

°3 missing; *categorized because of insufficient group discrimination

**S4 Table. Odds ratios of multivariate logistic regression for risk of unplanned 30-day readmission by social factors, health status and length of stay in hospital for ischaemic heart disease (N total=6,872/N readmissions=185)**

|  | **A: Social factors** | | | | **B: Health status** | | | | **C: Length of stay** | | | |
| --- | --- | --- | --- | --- | --- | --- | --- | --- | --- | --- | --- | --- |
|  | Sig. | Exp(B) | 95% CI | | Sig. | Exp(B) | 95% CI | | Sig. | Exp(B) | 95% CI | |
|  |  |  | Lower | Upper |  |  | Lower | Upper |  |  | Lower | Upper |
| Education level |  |  |  |  |  |  |  |  |  |  |  |  |
| tertiary (ref.) | 0.413 |  |  |  | 0.524 |  |  |  | 0.63 |  |  |  |
| upper secondary | 0.868 | 0.967 | 0.648 | 1.442 | 0.714 | 0.927 | 0.619 | 1.388 | 0.795 | 0.948 | 0.632 | 1.421 |
| compulsory | 0.387 | 1.222 | 0.776 | 1.923 | 0.583 | 1.137 | 0.718 | 1.801 | 0.61 | 1.128 | 0.71 | 1.792 |
| Insurance class |  |  |  |  |  |  |  |  |  |  |  |  |
| mandatory (ref.) |  |  |  |  |  |  |  |  |  |  |  |  |
| (Semi-) private | 0.35 | 0.857 | 0.62 | 1.185 | 0.631 | 0.922 | 0.663 | 1.283 | 0.774 | 0.953 | 0.683 | 1.328 |
| Household type |  |  |  |  |  |  |  |  |  |  |  |  |
| Living with others (ref.) |  |  |  |  |  |  |  |  |  |  |  |  |
| Living alone | 0.135 | 1.293 | 0.923 | 1.81 | 0.215 | 1.24 | 0.883 | 1.74 | 0.289 | 1.203 | 0.855 | 1.693 |
| Sex |  |  |  |  |  |  |  |  |  |  |  |  |
| Men (ref.) |  |  |  |  |  |  |  |  |  |  |  |  |
| Women | 0.122 | 1.304 | 0.931 | 1.827 | 0.074 | 1.364 | 0.971 | 1.917 | 0.082 | 1.355 | 0.963 | 1.908 |
| Age (years) | <.001 | 1.038 | 1.022 | 1.054 | <.001 | 1.03 | 1.014 | 1.046 | 0.001 | 1.026 | 1.01 | 1.042 |
| Comorbidity |  |  |  |  |  |  |  |  |  |  |  |  |
| Somatic Comorbidities: 0 (Ref.) |  |  |  |  | 0.04 |  |  |  | 0.333 |  |  |  |
| 1 |  |  |  |  | 0.689 | 1.096 | 0.698 | 1.721 | 0.813 | 1.056 | 0.672 | 1.661 |
| 2 |  |  |  |  | 0.048 | 1.575 | 1.004 | 2.469 | 0.157 | 1.389 | 0.882 | 2.189 |
| 3+ |  |  |  |  | 0.02 | 1.719 | 1.088 | 2.717 | 0.176 | 1.381 | 0.865 | 2.207 |
| Mental comorbidity: no (Ref.) |  |  |  |  |  |  |  |  |  |  |  |  |
| Mental comorbidity: yes |  |  |  |  | 0.023 | 1.993 | 1.098 | 3.619 | 0.195 | 1.496 | 0.813 | 2.752 |
| Previous hospital stay last 6 months |  |  |  |  |  |  |  |  |  |  |  |  |
| No (Ref.) |  |  |  |  |  |  |  |  |  |  |  |  |
| Yes |  |  |  |  | <.001 | 3.152 | 2.285 | 4.348 | <.001 | 3.18 | 2.301 | 4.395 |
| LOS, centred by CHC, Q1-Q3 (Ref.) |  |  |  |  |  |  |  |  |  |  |  |  |
| LOS, centred by CHC, Q4 |  |  |  |  |  |  |  |  | <.001 | 2.283 | 1.655 | 3.149 |
| Constant | <.001 | 0.002 |  |  | <.001 | 0.002 |  |  | <.001 | 0.002 |  |  |
| Omnibus Chi^2^ | 43.90(6). <.001 | | | | 103.65(11). <.001 | | | | 127.63(12). <.001 | | | |
| "-2 log-likelihood" | 1658.57 | | | | 1598.82 | | | | 1574.84 | | | |
| ROC | 0.645 | | | | 0.706 | | | | 0.731 | | | |

**S5 Table. Odds ratios of multivariate logistic regression for risk of unplanned 30-day readmission by social factors, health status and length of stay in hospital for congestive heart failure (N total=3,492°/N readmissions=324)**

|  | **A: Social factors** | | | | **B: Health status** | | | | **C: Length of stay** | | | |
| --- | --- | --- | --- | --- | --- | --- | --- | --- | --- | --- | --- | --- |
|  | Sig. | Exp(B) | 95% CI | | Sig. | Exp(B) | 95% CI | | Sig. | Exp(B) | 95% CI | |
|  |  |  | Lower | Upper |  |  | Lower | Upper |  |  | Lower | Upper |
| Education level |  |  |  |  |  |  |  |  |  |  |  |  |
| tertiary (ref.) | 0.042 |  |  |  | 0.048 |  |  |  | 0.057 |  |  |  |
| upper secondary | 0.118 | 1.37 | 0.923 | 2.034 | 0.134 | 1.354 | 0.911 | 2.011 | 0.132 | 1.357 | 0.912 | 2.017 |
| compulsory | 0.015 | 1.667 | 1.105 | 2.515 | 0.018 | 1.648 | 1.091 | 2.488 | 0.02 | 1.632 | 1.08 | 2.466 |
| Insurance class |  |  |  |  |  |  |  |  |  |  |  |  |
| mandatory (ref.) |  |  |  |  |  |  |  |  |  |  |  |  |
| (Semi-) private | 0.065 | 1.29 | 0.985 | 1.691 | 0.063 | 1.293 | 0.986 | 1.697 | 0.043 | 1.325 | 1.009 | 1.739 |
| Household type |  |  |  |  |  |  |  |  |  |  |  |  |
| Living with others (ref.) |  |  |  |  |  |  |  |  |  |  |  |  |
| Living alone | 0.653 | 1.06 | 0.823 | 1.364 | 0.553 | 1.08 | 0.838 | 1.392 | 0.604 | 1.069 | 0.83 | 1.379 |
| Sex |  |  |  |  |  |  |  |  |  |  |  |  |
| Men (ref.) |  |  |  |  |  |  |  |  |  |  |  |  |
| Women | 0.316 | 0.876 | 0.676 | 1.135 | 0.359 | 0.885 | 0.682 | 1.149 | 0.333 | 0.879 | 0.677 | 1.141 |
| Age (years) |  |  |  |  |  |  |  |  |  |  |  |  |
| <74 | 0.102 |  |  |  | 0.166 |  |  |  | 0.173 |  |  |  |
| 74-81 | 0.059 | 1.363 | 0.989 | 1.878 | 0.116 | 1.296 | 0.938 | 1.791 | 0.107 | 1.305 | 0.944 | 1.803 |
| 82-87 | 0.186 | 1.249 | 0.898 | 1.737 | 0.321 | 1.183 | 0.849 | 1.65 | 0.349 | 1.172 | 0.841 | 1.635 |
| 88+ | 0.019 | 1.529 | 1.073 | 2.178 | 0.031 | 1.477 | 1.035 | 2.106 | 0.036 | 1.462 | 1.025 | 2.087 |
| Comorbidity |  |  |  |  |  |  |  |  |  |  |  |  |
| Somatic Comorbidities: 0-2 (ref.) |  |  |  |  | 0.006 |  |  |  | 0.018 |  |  |  |
| 3 |  |  |  |  | 0.332 | 0.83 | 0.57 | 1.209 | 0.3 | 0.819 | 0.562 | 1.194 |
| 4 |  |  |  |  | 0.224 | 1.236 | 0.879 | 1.738 | 0.295 | 1.2 | 0.853 | 1.69 |
| 5+ |  |  |  |  | 0.018 | 1.458 | 1.068 | 1.989 | 0.045 | 1.377 | 1.007 | 1.883 |
| Mental comorbidity: no (ref.) |  |  |  |  |  |  |  |  |  |  |  |  |
| Mental comorbidity: yes |  |  |  |  | 0.9 | 1.022 | 0.73 | 1.43 | 0.998 | 1 | 0.714 | 1.402 |
| Previous hospital stay last 6 months |  |  |  |  |  |  |  |  |  |  |  |  |
| No (Ref.) |  |  |  |  |  |  |  |  |  |  |  |  |
| Yes |  |  |  |  | 0.175 | 1.19 | 0.926 | 1.531 | 0.187 | 1.185 | 0.921 | 1.525 |
| LOS, centred by CHC, Q1-Q3 (Ref.) |  |  |  |  |  |  |  |  |  |  |  |  |
| LOS, centred by CHC, Q4 |  |  |  |  |  |  |  |  | <.001 | 1.651 | 1.279 | 2.132 |
| Constant | <.001 | 0.055 |  |  | <.001 | 0.047 |  |  | <.001 | 0.043 |  |  |
| Omnibus Chi^2^ | 16.02(8) p. <.05 | | | | 31.31(13) p. <.01 | | | | 45.35(14) p. <.001 | | | |
| "-2 log-likelihood" | 2141.55 | | | | 2126.27 | | | | 2112.23 | | | |
| ROC | 0.563 | | | | 0.594 | | | | 0.614 | | | |

**°** 1 Missing: n=2

**S6 Table. Odds ratios of multivariate logistic regression for risk of unplanned 30-day readmission by social factors, health status and length of stay in hospital for acute myocard infarctus (N total=4,275/N readmissions=166)**

|  | **A: Social factors** | | | | **B: Health status** | | | | **C: Length of stay** | | | |
| --- | --- | --- | --- | --- | --- | --- | --- | --- | --- | --- | --- | --- |
|  | Sig. | Exp(B) | 95% CI | | Sig. | Exp(B) | 95% CI | | Sig. | Exp(B) | 95% CI | |
|  |  |  | Lower | Upper |  |  | Lower | Upper |  |  | Lower | Upper |
| Education level |  |  |  |  |  |  |  |  |  |  |  |  |
| tertiary (ref.) | 0.258 |  |  |  | 0.267 |  |  |  | 0.294 |  |  |  |
| upper secondary | 0.122 | 1.413 | 0.911 | 2.191 | 0.136 | 1.398 | 0.9 | 2.17 | 0.167 | 1.365 | 0.878 | 2.12 |
| compulsory | 0.512 | 1.188 | 0.71 | 1.986 | 0.572 | 1.16 | 0.693 | 1.944 | 0.669 | 1.119 | 0.668 | 1.876 |
| Insurance class |  |  |  |  |  |  |  |  |  |  |  |  |
| mandatory (ref.) |  |  |  |  |  |  |  |  |  |  |  |  |
| (Semi-) private | 0.197 | 0.776 | 0.528 | 1.141 | 0.204 | 0.779 | 0.53 | 1.146 | 0.234 | 0.791 | 0.538 | 1.164 |
| Household type |  |  |  |  |  |  |  |  |  |  |  |  |
| Living with others (ref.) |  |  |  |  |  |  |  |  |  |  |  |  |
| Living alone | 0.921 | 1.019 | 0.707 | 1.468 | 0.974 | 0.994 | 0.689 | 1.434 | 0.846 | 0.964 | 0.668 | 1.392 |
| Sex |  |  |  |  |  |  |  |  |  |  |  |  |
| Men (ref.) |  |  |  |  |  |  |  |  |  |  |  |  |
| Women | 0.475 | 1.145 | 0.79 | 1.661 | 0.389 | 1.178 | 0.812 | 1.71 | 0.456 | 1.152 | 0.794 | 1.672 |
| Age (years) | <.001 | 1.04 | 1.026 | 1.053 | <.001 | 1.033 | 1.019 | 1.047 | <.001 | 1.029 | 1.015 | 1.044 |
| Comorbidity |  |  |  |  |  |  |  |  |  |  |  |  |
| Somatic Comorbidities: 0 (ref.) |  |  |  |  | 0.013 |  |  |  | 0.111 |  |  |  |
| 1 |  |  |  |  | 0.703 | 1.095 | 0.687 | 1.744 | 0.826 | 1.054 | 0.661 | 1.681 |
| 2 |  |  |  |  | 0.697 | 0.899 | 0.526 | 1.536 | 0.468 | 0.819 | 0.477 | 1.405 |
| 3+ |  |  |  |  | 0.018 | 1.773 | 1.103 | 2.852 | 0.14 | 1.449 | 0.885 | 2.371 |
| Mental comorbidity: no (ref.) |  |  |  |  |  |  |  |  |  |  |  |  |
| Mental comorbidity: yes |  |  |  |  | 0.216 | 0.671 | 0.357 | 1.261 | 0.117 | 0.602 | 0.319 | 1.135 |
| Previous hospital stay last 6 months |  |  |  |  |  |  |  |  |  |  |  |  |
| no (ref.) |  |  |  |  |  |  |  |  |  |  |  |  |
| yes |  |  |  |  | 0.043 | 1.652 | 1.015 | 2.687 | 0.054 | 1.614 | 0.991 | 2.629 |
| LOS, centred by CHC, Q1-Q3 (ref.) |  |  |  |  |  |  |  |  |  |  |  |  |
| LOS, centred by CHC, Q4 |  |  |  |  |  |  |  |  | 0.001 | 1.819 | 1.283 | 2.578 |
| Constant | <.001 | 0.002 |  |  | <.001 | 0.003 |  |  | <.001 | 0.004 |  |  |
| Omnibus Chi^2^ | 44.21(6) p<.001 | | | | 60.81(11) p<.001 | | | | 71.84(12) p<.001 | | | |
| "-2 log-likelihood" | 1359.78 | | | | 1343.18 | | | | 1332.15 | | | |
| ROC | 0.648 | | | | 0.665 | | | | 0.681 | | | |

**S7 Table. Odds ratios of multivariate logistic regression for risk of unplanned 30-day readmission by social factors, health status and length of stay in hospital for acute cerebrovascular disease (stroke) (N total=3,109/N readmissions=143)**

|  | **A: Social factors** | | | | **B: Health status** | | | | **C: Length of stay** | | | |
| --- | --- | --- | --- | --- | --- | --- | --- | --- | --- | --- | --- | --- |
|  | Sig. | Exp(B) | 95% CI | | Sig. | Exp(B) | 95% CI | | Sig. | Exp(B) | 95% CI | |
|  |  |  | Lower | Upper |  |  | Lower | Upper |  |  | Lower | Upper |
| Education level |  |  |  |  |  |  |  |  |  |  |  |  |
| tertiary (ref.) | 0.143 |  |  |  | 0.156 |  |  |  | 0.163 |  |  |  |
| upper secondary | 0.42 | 1.232 | 0.742 | 2.045 | 0.425 | 1.233 | 0.737 | 2.064 | 0.421 | 1.235 | 0.738 | 2.068 |
| compulsory | 0.07 | 1.658 | 0.959 | 2.869 | 0.076 | 1.652 | 0.948 | 2.877 | 0.079 | 1.645 | 0.944 | 2.867 |
| Insurance class |  |  |  |  |  |  |  |  |  |  |  |  |
| mandatory (ref.) |  |  |  |  |  |  |  |  |  |  |  |  |
| (Semi-)private | 0.536 | 0.879 | 0.583 | 1.324 | 0.578 | 0.888 | 0.584 | 1.35 | 0.584 | 0.89 | 0.585 | 1.353 |
| Household type |  |  |  |  |  |  |  |  |  |  |  |  |
| Living with others (ref.) |  |  |  |  |  |  |  |  |  |  |  |  |
| Living alone | 0.333 | 1.206 | 0.826 | 1.76 | 0.443 | 1.161 | 0.792 | 1.702 | 0.469 | 1.152 | 0.786 | 1.689 |
| Sex |  |  |  |  |  |  |  |  |  |  |  |  |
| Men (ref.) |  |  |  |  |  |  |  |  |  |  |  |  |
| Women | 0.37 | 0.841 | 0.575 | 1.229 | 0.386 | 0.844 | 0.576 | 1.237 | 0.358 | 0.836 | 0.57 | 1.226 |
| Age (years) | <.001 | 1.03 | 1.015 | 1.045 | 0.007 | 1.021 | 1.006 | 1.037 | 0.01 | 1.02 | 1.005 | 1.036 |
| Comorbidity |  |  |  |  |  |  |  |  |  |  |  |  |
| Somatic Comorbidities: 0 (ref.) |  |  |  |  | 0.013 |  |  |  | 0.015 |  |  |  |
| 1 |  |  |  |  | 0.245 | 1.525 | 0.749 | 3.106 | 0.218 | 1.567 | 0.767 | 3.2 |
| 2 |  |  |  |  | 0.265 | 1.495 | 0.737 | 3.034 | 0.247 | 1.52 | 0.748 | 3.089 |
| 3+ |  |  |  |  | 0.009 | 2.46 | 1.257 | 4.814 | 0.008 | 2.477 | 1.265 | 4.848 |
| Mental comorbidity: no (ref.) |  |  |  |  |  |  |  |  |  |  |  |  |
| Mental comorbidity: yes |  |  |  |  | 0.022 | 1.672 | 1.077 | 2.596 | 0.027 | 1.646 | 1.059 | 2.559 |
| Previous hospital stay last 6 months |  |  |  |  |  |  |  |  |  |  |  |  |
| No (ref.) |  |  |  |  |  |  |  |  |  |  |  |  |
| Yes |  |  |  |  | <.001 | 3.676 | 2.48 | 5.449 | <.001 | 3.698 | 2.494 | 5.483 |
| LOS, centred by CHC, Q1-Q3 (Ref.) |  |  |  |  |  |  |  |  |  |  |  |  |
| LOS, centred by CHC, Q4 |  |  |  |  |  |  |  |  | 0.399 | 1.185 | 0.799 | 1.758 |
| Constant | <.001 | 0.005 |  |  | <.001 | 0.004 |  |  | <.001 | 0.004 |  |  |
| Omnibus Chi^2^ | 28.75(6) p<.001 | | | | 83.04(11) p<.001 | | | | 84.74(12) p<.001 | | | |
| "-2 log-likelihood" | 1131.22 | | | | 1076.93 | | | | 1076.24 | | | |
| ROC | 0.63 | | | | 0.72 | | | | 0.721 | | | |

**S8 Table. Odds ratios of multivariate logistic regression for risk of unplanned 30-day readmission by social factors, health status and length of stay in hospital for all cancer (N total=10,165/N readmissions=516)**

|  | **A: Social factors** | | | | **B: Health status** | | | | **C: Length of stay** | | | |
| --- | --- | --- | --- | --- | --- | --- | --- | --- | --- | --- | --- | --- |
|  | Sig. | Exp(B) | 95% CI | | Sig. | Exp(B) | 95% CI | | Sig. | Exp(B) | 95% CI | |
|  |  |  | Lower | Upper |  |  | Lower | Upper |  |  | Lower | Upper |
| Education level |  |  |  |  |  |  |  |  |  |  |  |  |
| tertiary (ref.) | <.001 |  |  |  | 0.001 |  |  |  | 0.001 |  |  |  |
| upper secondary | 0.02 | 1.35 | 1.049 | 1.736 | 0.038 | 1.306 | 1.015 | 1.682 | 0.034 | 1.314 | 1.02 | 1.692 |
| compulsory | <.001 | 1.825 | 1.375 | 2.422 | <.001 | 1.709 | 1.286 | 2.272 | <.001 | 1.702 | 1.28 | 2.263 |
| Insurance class |  |  |  |  |  |  |  |  |  |  |  |  |
| mandatory (ref.) |  |  |  |  |  |  |  |  |  |  |  |  |
| (Semi-)private | 0.077 | 0.832 | 0.678 | 1.02 | 0.156 | 0.862 | 0.702 | 1.058 | 0.161 | 0.863 | 0.703 | 1.06 |
| Household type |  |  |  |  |  |  |  |  |  |  |  |  |
| Living with others (ref.) |  |  |  |  |  |  |  |  |  |  |  |  |
| Living alone | 0.194 | 1.149 | 0.932 | 1.416 | 0.274 | 1.124 | 0.911 | 1.387 | 0.373 | 1.101 | 0.892 | 1.359 |
| Sex |  |  |  |  |  |  |  |  |  |  |  |  |
| Men (Ref.) |  |  |  |  |  |  |  |  |  |  |  |  |
| Women | <.001 | 0.47 | 0.387 | 0.571 | <.001 | 0.475 | 0.391 | 0.577 | <.001 | 0.466 | 0.383 | 0.567 |
| Age (years) | 0.005 | 1.012 | 1.004 | 1.021 | 0.184 | 1.006 | 0.997 | 1.015 | 0.199 | 1.006 | 0.997 | 1.014 |
| Comorbidity |  |  |  |  |  |  |  |  |  |  |  |  |
| NSD, centered by CHC |  |  |  |  | <.001 | 1.193 | 1.111 | 1.281 | <.001 | 1.155 | 1.074 | 1.242 |
| Mental comorbidity: no (ref.) |  |  |  |  |  |  |  |  |  |  |  |  |
| Mental comorbidity: yes |  |  |  |  | 0.006 | 1.576 | 1.137 | 2.185 | 0.011 | 1.526 | 1.1 | 2.117 |
| Previous hospital stay last 6 months |  |  |  |  |  |  |  |  |  |  |  |  |
| No (ref.) |  |  |  |  |  |  |  |  |  |  |  |  |
| Yes |  |  |  |  | 0.001 | 1.474 | 1.17 | 1.858 | 0.001 | 1.467 | 1.163 | 1.849 |
| LOS, centred by CHC, Q1-Q3 (Ref.) |  |  |  |  |  |  |  |  |  |  |  |  |
| LOS, centred by CHC, Q4 |  |  |  |  |  |  |  |  | <.001 | 1.551 | 1.268 | 1.896 |
| Constant | <.001 | 0.025 |  |  | <.001 | 0.035 |  |  | <.001 | 0.032 |  |  |
| Omnibus Chi^2^ | 111.52(6) p<.001 | | | | 155.82(9) p<.001 | | | | 173.27(10) p<.001 | | | |
| "-2 log-likelihood" | 3969.82 | | | | 3925.51 | | | | 3908.06 | | | |
| ROC | 0.635 | | | | 0.663 | | | | 0.671 | | | |

**S9 Table. Odds ratios of multivariate logistic regression for risk of unplanned 30-day readmission by social factors, health status and length of stay in hospital for lung cancer (N total=1'418/N readmissions=200)**

|  | **A: Social factors** | | | | **B: Health status** | | | | **C: Length of stay** | | | |
| --- | --- | --- | --- | --- | --- | --- | --- | --- | --- | --- | --- | --- |
|  | Sig. | Exp(B) | 95% CI | | Sig. | Exp(B) | 95% CI | | Sig. | Exp(B) | 95% CI | |
|  |  |  | Lower | Upper |  |  | Lower | Upper |  |  | Lower | Upper |
| Education level |  |  |  |  |  |  |  |  |  |  |  |  |
| tertiary (ref.) | 0.007 |  |  |  | 0.01 |  |  |  | 0.007 |  |  |  |
| upper secondary | 0.008 | 2.019 | 1.198 | 3.401 | 0.011 | 1.972 | 1.168 | 3.329 | 0.007 | 2.07 | 1.223 | 3.503 |
| compulsory | 0.002 | 2.418 | 1.39 | 4.204 | 0.002 | 2.359 | 1.354 | 4.109 | 0.002 | 2.42 | 1.387 | 4.22 |
| Insurance class |  |  |  |  |  |  |  |  |  |  |  |  |
| mandatory (ref.) |  |  |  |  |  |  |  |  |  |  |  |  |
| (Semi-) private | 0.886 | 1.027 | 0.712 | 1.482 | 0.684 | 1.079 | 0.747 | 1.561 | 0.681 | 1.081 | 0.746 | 1.565 |
| Household type |  |  |  |  |  |  |  |  |  |  |  |  |
| Living with others (ref.) |  |  |  |  |  |  |  |  |  |  |  |  |
| Living alone | 0.672 | 0.928 | 0.658 | 1.31 | 0.618 | 0.915 | 0.646 | 1.296 | 0.542 | 0.897 | 0.632 | 1.272 |
| Sex |  |  |  |  |  |  |  |  |  |  |  |  |
| Men (ref.) |  |  |  |  |  |  |  |  |  |  |  |  |
| Women | 0.154 | 0.79 | 0.571 | 1.093 | 0.22 | 0.814 | 0.587 | 1.13 | 0.206 | 0.809 | 0.582 | 1.124 |
| Age (years) | 0.522 | 1.005 | 0.99 | 1.02 | 0.88 | 0.999 | 0.983 | 1.015 | 0.812 | 0.998 | 0.982 | 1.014 |
| Comorbidity |  |  |  |  |  |  |  |  |  |  |  |  |
| Somatic Comorbidities: 0 (ref.) |  |  |  |  | 0.003 |  |  |  | 0.008 |  |  |  |
| 1 |  |  |  |  | 0.026 | 1.933 | 1.083 | 3.451 | 0.029 | 1.906 | 1.067 | 3.406 |
| 2 |  |  |  |  | 0.027 | 1.974 | 1.081 | 3.604 | 0.055 | 1.81 | 0.987 | 3.319 |
| 3+ |  |  |  |  | <.001 | 2.895 | 1.625 | 5.159 | 0.001 | 2.656 | 1.485 | 4.749 |
| Mental comorbidity: no (ref.) |  |  |  |  |  |  |  |  |  |  |  |  |
| Mental comorbidity: yes |  |  |  |  | 0.936 | 0.98 | 0.589 | 1.628 | 0.87 | 0.958 | 0.576 | 1.595 |
| Previous hospital stay last 6 months |  |  |  |  |  |  |  |  |  |  |  |  |
| No (ref.) |  |  |  |  |  |  |  |  |  |  |  |  |
| Yes |  |  |  |  | 0.401 | 1.164 | 0.816 | 1.659 | 0.444 | 1.149 | 0.805 | 1.64 |
| LOS, centred by CHC, Q1-Q3 (Ref.) |  |  |  |  |  |  |  |  |  |  |  |  |
| LOS, centred by CHC, Q4 |  |  |  |  |  |  |  |  | 0.005 | 1.636 | 1.165 | 2.299 |
| Constant | <.001 | 0.067 |  |  | <.001 | 0.047 |  |  | <.001 | 0.045 |  |  |
| Omnibus Chi^2^ | 13.93(6) p<.05 | | | | 30.03(11) p<.01 | | | | 37.79(12) p<.001 | | | |
| "-2 log-likelihood" | 1139.91 | | | | 1123.81 | | | | 1116.05 | | | |
| ROC | 0.568 | | | | 0.624 | | | | 0.635 | | | |

**S10 Table. Odds ratios of multivariate logistic regression for risk of unplanned 30-day readmission by social factors, health status and length of stay in hospital for breast cancer (N total=4'547/N readmissions=101)**

|  | **A: Social factors** | | | | **B: Health status** | | | | **C: Length of stay** | | | |
| --- | --- | --- | --- | --- | --- | --- | --- | --- | --- | --- | --- | --- |
|  | Sig. | Exp(B) | 95% CI | | Sig. | Exp(B) | 95% CI | | Sig. | Exp(B) | 95% CI | |
|  |  |  | Lower | Upper |  |  | Lower | Upper |  |  | Lower | Upper |
| Education level |  |  |  |  |  |  |  |  |  |  |  |  |
| tertiary (ref.) | 0.31 |  |  |  | 0.537 |  |  |  | 0.531 |  |  |  |
| upper secondary | 0.576 | 1.183 | 0.656 | 2.136 | 0.694 | 1.127 | 0.622 | 2.04 | 0.678 | 1.134 | 0.626 | 2.055 |
| compulsory | 0.168 | 1.577 | 0.826 | 3.013 | 0.318 | 1.395 | 0.725 | 2.684 | 0.31 | 1.405 | 0.729 | 2.707 |
| Insurance class |  |  |  |  |  |  |  |  |  |  |  |  |
| mandatory (ref.) |  |  |  |  |  |  |  |  |  |  |  |  |
| (Semi-)private | 0.666 | 0.91 | 0.592 | 1.398 | 0.908 | 0.975 | 0.632 | 1.504 | 0.909 | 0.975 | 0.632 | 1.505 |
| Household type |  |  |  |  |  |  |  |  |  |  |  |  |
| Living with others (ref.) |  |  |  |  |  |  |  |  |  |  |  |  |
| Living alone | 0.581 | 1.133 | 0.728 | 1.764 | 0.619 | 1.119 | 0.718 | 1.744 | 0.743 | 1.078 | 0.689 | 1.685 |
| Age (years) | 0.276 | 1.009 | 0.992 | 1.027 | 0.996 | 1 | 0.983 | 1.018 | 0.974 | 1 | 0.982 | 1.017 |
| Comorbidity |  |  |  |  |  |  |  |  |  |  |  |  |
| Somatic Comorbidities: 0 (ref.) |  |  |  |  | 0.003 |  |  |  | 0.023 |  |  |  |
| 1 |  |  |  |  | 0.108 | 1.483 | 0.917 | 2.398 | 0.197 | 1.378 | 0.846 | 2.243 |
| 2 |  |  |  |  | 0.001 | 2.577 | 1.439 | 4.615 | 0.007 | 2.281 | 1.255 | 4.145 |
| 3+ |  |  |  |  | 0.004 | 2.837 | 1.398 | 5.757 | 0.017 | 2.422 | 1.17 | 5.012 |
| Mental comorbidity: no (ref.) |  |  |  |  |  |  |  |  |  |  |  |  |
| Mental comorbidity: yes |  |  |  |  | 0.072 | 1.872 | 0.945 | 3.706 | 0.078 | 1.849 | 0.934 | 3.661 |
| Previous hospital stay last 6 months |  |  |  |  |  |  |  |  |  |  |  |  |
| No (ref.) |  |  |  |  |  |  |  |  |  |  |  |  |
| Yes |  |  |  |  | 0.178 | 1.511 | 0.829 | 2.754 | 0.205 | 1.475 | 0.809 | 2.69 |
| LOS, centred by CHC, Q1-Q3 (ref.) |  |  |  |  |  |  |  |  |  |  |  |  |
| LOS, centred by CHC, Q4 |  |  |  |  |  |  |  |  | 0.042 | 1.563 | 1.015 | 2.407 |
| Constant | <.001 | 0.01 |  |  | <.001 | 0.012 |  |  | <.001 | 0.012 |  |  |
| Omnibus Chi^2^ | 6.77(5) p=.238 | | | | 26.90(10) p<.01 | | | | 30.88(11) p=.001 | | | |
| "-2 log-likelihood" | 962 | | | | 941.88 | | | | 937.9 | | | |
| ROC | 0.574 | | | | 0.635 | | | | 0.637 | | | |

**S11 Table. Odds ratios of multivariate logistic regression for risk of unplanned 30-day readmission by social factors, health status and length of stay in hospital for prostate cancer (N total=2,806/N readmissions=145)**

|  | **A: Social factors** | | | | **B: Health status** | | | | **C: length of stay** | | | |
| --- | --- | --- | --- | --- | --- | --- | --- | --- | --- | --- | --- | --- |
|  | Sig. | Exp(B) | 95% CI | | Sig. | Exp(B) | 95% CI | | Sig. | Exp(B) | 95% CI | |
|  |  |  | Lower | Upper |  |  | Lower | Upper |  |  | Lower | Upper |
| Education level |  |  |  |  |  |  |  |  |  |  |  |  |
| tertiary (ref.) | 0.443 |  |  |  | 0.479 |  |  |  | 0.515 |  |  |  |
| upper secondary | 0.349 | 0.827 | 0.557 | 1.23 | 0.29 | 0.807 | 0.542 | 1.201 | 0.296 | 0.809 | 0.544 | 1.204 |
| compulsory | 0.807 | 1.064 | 0.645 | 1.758 | 0.958 | 0.986 | 0.594 | 1.637 | 0.893 | 0.966 | 0.581 | 1.605 |
| Insurance class |  |  |  |  |  |  |  |  |  |  |  |  |
| mandatory (ref.) |  |  |  |  |  |  |  |  |  |  |  |  |
| (Semi-) private | 0.402 | 0.852 | 0.586 | 1.239 | 0.516 | 0.883 | 0.606 | 1.286 | 0.499 | 0.878 | 0.603 | 1.28 |
| Household type |  |  |  |  |  |  |  |  |  |  |  |  |
| Living with others (ref.) |  |  |  |  |  |  |  |  |  |  |  |  |
| Living alone | 0.296 | 1.252 | 0.821 | 1.909 | 0.319 | 1.241 | 0.812 | 1.896 | 0.325 | 1.237 | 0.809 | 1.892 |
| Age (years) | 0.149 | 1.014 | 0.995 | 1.033 | 0.775 | 1.003 | 0.983 | 1.023 | 0.747 | 1.003 | 0.984 | 1.023 |
| Comorbidity |  |  |  |  |  |  |  |  |  |  |  |  |
| Somatic Comorbidities: 0 (ref.) |  |  |  |  | 0.027 |  |  |  | 0.067 |  |  |  |
| 1 |  |  |  |  | 0.182 | 1.316 | 0.88 | 1.968 | 0.2 | 1.302 | 0.87 | 1.949 |
| 2 |  |  |  |  | 0.249 | 1.358 | 0.807 | 2.286 | 0.318 | 1.305 | 0.774 | 2.202 |
| 3+ |  |  |  |  | 0.003 | 2.43 | 1.366 | 4.322 | 0.008 | 2.21 | 1.231 | 3.969 |
| Mental comorbidity: no (ref.) |  |  |  |  |  |  |  |  |  |  |  |  |
| Mental comorbidity: yes |  |  |  |  | 0.277 | 1.492 | 0.725 | 3.074 | 0.336 | 1.427 | 0.692 | 2.945 |
| Previous hospital stay last 6 months |  |  |  |  |  |  |  |  |  |  |  |  |
| no (ref.) |  |  |  |  |  |  |  |  |  |  |  |  |
| yes |  |  |  |  | 0.144 | 1.394 | 0.893 | 2.175 | 0.17 | 1.366 | 0.875 | 2.135 |
| LOS, centred by CHC, Q1-Q3 (Ref.) |  |  |  |  |  |  |  |  |  |  |  |  |
| LOS, centred by CHC, Q4 |  |  |  |  |  |  |  |  | 0.039 | 1.523 | 1.022 | 2.27 |
| Constant | <.001 | 0.023 |  |  | <.001 | 0.038 |  |  | <.001 | 0.035 |  |  |
| Omnibus Chi^2^ | 5.84(5) p=.322 | | | | 19.31(10) p<.05 | | | | 23.34(11) p<.05 | | | |
| "-2 log-likelihood" | 1135.74 | | | | 1122.28 | | | | 1118.24 | | | |
| ROC | 0.558 | | | | 0.608 | | | | 0.616 | | | |

**S12 Table. Odds ratios of multivariate logistic regression for risk of unplanned 30-day readmission by social factors, health status and length of stay in hospital for COPD (N total=2115/N readmissions=129)**

|  | **A: Social factors** | | | | **B: Health status** | | | | **C: Length of stay** | | | |
| --- | --- | --- | --- | --- | --- | --- | --- | --- | --- | --- | --- | --- |
|  | Sig. | Exp(B) | 95% CI | | Sig. | Exp(B) | 95% CI | | Sig. | Exp(B) | 95% CI | |
|  |  |  | Lower | Upper |  |  | Lower | Upper |  |  | Lower | Upper |
| Education level |  |  |  |  |  |  |  |  |  |  |  |  |
| tertiary (ref.) | 0.081 |  |  |  | 0.102 |  |  |  | 0.105 |  |  |  |
| upper secondary | 0.035 | 2.352 | 1.062 | 5.211 | 0.045 | 2.265 | 1.019 | 5.035 | 0.047 | 2.247 | 1.011 | 4.996 |
| compulsory | 0.127 | 1.889 | 0.834 | 4.279 | 0.147 | 1.837 | 0.808 | 4.18 | 0.154 | 1.818 | 0.799 | 4.133 |
| Insurance class |  |  |  |  |  |  |  |  |  |  |  |  |
| mandatory (ref.) |  |  |  |  |  |  |  |  |  |  |  |  |
| (Semi-) private | 0.222 | 0.747 | 0.467 | 1.193 | 0.181 | 0.725 | 0.452 | 1.162 | 0.176 | 0.722 | 0.45 | 1.157 |
| Household type |  |  |  |  |  |  |  |  |  |  |  |  |
| Living with others (ref.) |  |  |  |  |  |  |  |  |  |  |  |  |
| Living alone | 0.685 | 0.922 | 0.624 | 1.363 | 0.541 | 0.884 | 0.595 | 1.312 | 0.535 | 0.883 | 0.595 | 1.31 |
| Sex |  |  |  |  |  |  |  |  |  |  |  |  |
| Men (ref.) |  |  |  |  |  |  |  |  |  |  |  |  |
| Women | 0.604 | 0.903 | 0.614 | 1.328 | 0.835 | 0.959 | 0.649 | 1.419 | 0.825 | 0.957 | 0.648 | 1.414 |
| Age (years) | 0.014 | 1.022 | 1.004 | 1.04 | 0.074 | 1.017 | 0.998 | 1.036 | 0.09 | 1.016 | 0.998 | 1.035 |
| Comorbidity |  |  |  |  |  |  |  |  |  |  |  |  |
| Somatic Comorbidities: 0 (ref.) |  |  |  |  | 0.003 |  |  |  | 0.005 |  |  |  |
| 1 |  |  |  |  | 0.593 | 0.857 | 0.487 | 1.508 | 0.562 | 0.846 | 0.481 | 1.489 |
| 2 |  |  |  |  | 0.374 | 0.758 | 0.412 | 1.395 | 0.348 | 0.747 | 0.406 | 1.374 |
| 3+ |  |  |  |  | 0.037 | 1.724 | 1.034 | 2.876 | 0.055 | 1.656 | 0.988 | 2.774 |
| Mental comorbidity: no (ref.) |  |  |  |  |  |  |  |  |  |  |  |  |
| Mental comorbidity: yes |  |  |  |  | 0.411 | 0.808 | 0.486 | 1.343 | 0.379 | 0.796 | 0.478 | 1.324 |
| Previous hospital stay last 6 months |  |  |  |  |  |  |  |  |  |  |  |  |
| No (ref.) |  |  |  |  |  |  |  |  |  |  |  |  |
| Yes |  |  |  |  | 0.044 | 1.526 | 1.012 | 2.303 | 0.045 | 1.524 | 1.01 | 2.299 |
| LOS, centred by CHC, Q1-Q3 (Ref.) |  |  |  |  |  |  |  |  |  |  |  |  |
| LOS, centred by CHC, Q4 |  |  |  |  |  |  |  |  | 0.222 | 1.284 | 0.86 | 1.918 |
| Constant | <.001 | 0.008 |  |  | <.001 | 0.01 |  |  | <.001 | 0.01 |  |  |
| Omnibus Chi^2^ | 13.63(6) p<.05 | | | | 31.61(11) p<.001 | | | | 33.06(12) p<.001 | | | |
| "-2 log-likelihood" | 957.97 | | | | 939.99 | | | | 938.54 | | | |
| ROC | 0.587 | | | | 0.645 | | | | 0.640 | | | |

**S13 Table. Odds ratios of multivariate logistic regression for risk of unplanned 30-day readmission by social factors, health status and length of stay in hospital for back problems (N total=12,691/N readmissions=332)**

|  | **A: Social factors** | | | | **B: Health status** | | | | **C: Length of stay** | | | |
| --- | --- | --- | --- | --- | --- | --- | --- | --- | --- | --- | --- | --- |
|  | Sig. | Exp(B) | 95% CI | | Sig. | Exp(B) | 95% CI | | Sig. | Exp(B) | 95% CI | |
|  |  |  | Lower | Upper |  |  | Lower | Upper |  |  | Lower | Upper |
| Education level |  |  |  |  |  |  |  |  |  |  |  |  |
| tertiary (ref.) | 0.02 |  |  |  | 0.059 |  |  |  | 0.063 |  |  |  |
| upper secondary | 0.01 | 1.584 | 1.118 | 2.243 | 0.023 | 1.501 | 1.059 | 2.128 | 0.023 | 1.497 | 1.056 | 2.123 |
| compulsory | 0.007 | 1.694 | 1.152 | 2.491 | 0.028 | 1.542 | 1.047 | 2.272 | 0.032 | 1.527 | 1.037 | 2.25 |
| Insurance class |  |  |  |  |  |  |  |  |  |  |  |  |
| mandatory (ref.) |  |  |  |  |  |  |  |  |  |  |  |  |
| (Semi-) private | 0.016 | 0.724 | 0.557 | 0.942 | 0.03 | 0.746 | 0.572 | 0.972 | 0.022 | 0.734 | 0.563 | 0.956 |
| Household type |  |  |  |  |  |  |  |  |  |  |  |  |
| Living with others (ref.) |  |  |  |  |  |  |  |  |  |  |  |  |
| Living alone | 0.541 | 1.082 | 0.84 | 1.395 | 0.877 | 1.02 | 0.79 | 1.318 | 0.996 | 0.999 | 0.773 | 1.292 |
| Sex |  |  |  |  |  |  |  |  |  |  |  |  |
| Men (ref.) |  |  |  |  |  |  |  |  |  |  |  |  |
| Women | 0.554 | 0.932 | 0.737 | 1.178 | 0.574 | 0.935 | 0.739 | 1.183 | 0.494 | 0.921 | 0.728 | 1.166 |
| Age (years) | <.001 | 1.031 | 1.023 | 1.039 | <.001 | 1.02 | 1.012 | 1.029 | <.001 | 1.019 | 1.01 | 1.028 |
| Comorbidity |  |  |  |  |  |  |  |  |  |  |  |  |
| Somatic Comorbidities: 0 (ref.) |  |  |  |  |  |  |  |  |  |  |  |  |
| 1 |  |  |  |  | 0.001 | 1.616 | 1.222 | 2.138 | 0.001 | 1.605 | 1.213 | 2.123 |
| 2 |  |  |  |  | 0 | 1.89 | 1.355 | 2.636 | <.001 | 1.839 | 1.317 | 2.568 |
| 3+ |  |  |  |  | 0.001 | 1.898 | 1.281 | 2.813 | 0.003 | 1.83 | 1.234 | 2.716 |
| Mental comorbidity: no (ref.) |  |  |  |  |  |  |  |  |  |  |  |  |
| Mental comorbidity: yes |  |  |  |  | 0.008 | 1.544 | 1.118 | 2.132 | 0.023 | 1.46 | 1.054 | 2.022 |
| Previous hospital stay last 6 months |  |  |  |  |  |  |  |  |  |  |  |  |
| No (ref.) |  |  |  |  |  |  |  |  |  |  |  |  |
| Yes |  |  |  |  | 0 | 1.989 | 1.495 | 2.648 | <.001 | 1.961 | 1.473 | 2.611 |
| LOS, centred by CHC, Q1-Q3 (Ref.) |  |  |  |  |  |  |  |  |  |  |  |  |
| LOS, centred by CHC, Q4 |  |  |  |  |  |  |  |  | 0.011 | 1.389 | 1.079 | 1.788 |
| Constant | <.001 | 0.003 |  |  | <.001 | 0.004 |  |  | <.001 | 0.004 |  |  |
| Omnibus Chi^2^ | 88.32(6) p<.001 | | | | 142.80(11) p<.001 | | | | 149.08(12) p<.001 | | | |
| "-2 log-likelihood" | 2986.15 | | | | 2931.68 | | | | 2925.39 | | | |
| ROC | 0.645 | | | | 0.684 | | | | 0.690 | | | |

**S14 Table. Odds ratios of multivariate logistic regression for risk of unplanned 30-day readmission by social factors, health status and length of stay in hospital for osteoarthritis (N total=17,380/N readmissions=229)**

|  | **A: Social factors** | | | | **B: Health status** | | | | **C: Length of stay** | | | |
| --- | --- | --- | --- | --- | --- | --- | --- | --- | --- | --- | --- | --- |
|  | Sig. | Exp(B) | 95% CI | | Sig. | Exp(B) | 95% CI | | Sig. | Exp(B) | 95% CI | |
|  |  |  | Lower | Upper |  |  | Lower | Upper |  |  | Lower | Upper |
| Education level |  |  |  |  |  |  |  |  |  |  |  |  |
| tertiary (ref.) | 0.348 |  |  |  | 0.272 |  |  |  | 0.268 |  |  |  |
| upper secondary | 0.172 | 0.789 | 0.562 | 1.108 | 0.112 | 0.758 | 0.539 | 1.066 | 0.108 | 0.756 | 0.538 | 1.063 |
| compulsory | 0.627 | 0.905 | 0.605 | 1.353 | 0.424 | 0.848 | 0.565 | 1.271 | 0.395 | 0.839 | 0.559 | 1.258 |
| Insurance class |  |  |  |  |  |  |  |  |  |  |  |  |
| mandatory (ref.) |  |  |  |  |  |  |  |  |  |  |  |  |
| (Semi-) private | 0.396 | 0.883 | 0.662 | 1.177 | 0.432 | 0.89 | 0.667 | 1.189 | 0.298 | 0.857 | 0.64 | 1.147 |
| Household type |  |  |  |  |  |  |  |  |  |  |  |  |
| Living with others (ref.) |  |  |  |  |  |  |  |  |  |  |  |  |
| Living alone | 0.647 | 1.077 | 0.785 | 1.477 | 0.758 | 1.051 | 0.765 | 1.444 | 0.893 | 1.022 | 0.744 | 1.404 |
| Sex |  |  |  |  |  |  |  |  |  |  |  |  |
| Men (ref.) |  |  |  |  |  |  |  |  |  |  |  |  |
| Women | 0.011 | 0.692 | 0.521 | 0.919 | 0.015 | 0.702 | 0.527 | 0.934 | 0.011 | 0.691 | 0.519 | 0.92 |
| Age (years) | <.001 | 1.05 | 1.036 | 1.064 | <.001 | 1.043 | 1.029 | 1.057 | <.001 | 1.04 | 1.025 | 1.054 |
| Comorbidity |  |  |  |  |  |  |  |  |  |  |  |  |
| Somatic Comorbidities: 0 (ref.) |  |  |  |  | 0.022 |  |  |  | 0.058 |  |  |  |
| 1 |  |  |  |  | 0.151 | 1.263 | 0.918 | 1.737 | 0.179 | 1.244 | 0.904 | 1.712 |
| 2 |  |  |  |  | 0.007 | 1.67 | 1.152 | 2.42 | 0.015 | 1.587 | 1.093 | 2.304 |
| 3+ |  |  |  |  | 0.025 | 1.749 | 1.073 | 2.851 | 0.055 | 1.616 | 0.989 | 2.641 |
| Mental comorbidity: no (ref.) |  |  |  |  |  |  |  |  |  |  |  |  |
| Mental comorbidity: yes |  |  |  |  | 0.11 | 1.537 | 0.908 | 2.601 | 0.156 | 1.465 | 0.864 | 2.485 |
| Previous hospital stay last 6 months |  |  |  |  |  |  |  |  |  |  |  |  |
| No (ref.) |  |  |  |  |  |  |  |  |  |  |  |  |
| Yes |  |  |  |  | <.001 | 2.437 | 1.703 | 3.488 | <.001 | 2.444 | 1.708 | 3.499 |
| LOS, centred by CHC, Q1-Q3 (Ref.) |  |  |  |  |  |  |  |  |  |  |  |  |
| LOS, centred by CHC, Q4 |  |  |  |  |  |  |  |  | 0.002 | 1.548 | 1.17 | 2.048 |
| Constant | <.001 | 0.001 |  |  | <.001 | 0.001 |  |  | <.001 | 0.001 |  |  |
| Omnibus Chi^2^ | 67.22(6) p<.001 | | | | 102.23(11) p<.001 | | | | 111.28(12) p<.001 | | | |
| "-2 log-likelihood" | 2370.6 | | | | 2335.59 | | | | 2326.53 | | | |
| ROC | 0.647 | | | | 0.67 | | | | 0.681 | | | |
